# Effect of COVID-19 lockdown on sleep quality, sleep duration, and acute stress in Indian young adults

**DOI:** 10.1101/2025.08.15.25333638

**Authors:** Rishabh Soni, Ishan Gupta, Shifa Qureshi, Nasreen Akhtar

## Abstract

The COVID-19 lockdown brought disruptions to daily life and social schedules which influenced sleep and stress. Young adults, already vulnerable to irregular sleep patterns and heightened psychological stress, may have experienced notable shifts in sleep quality, duration, and circadian alignment during this period.

Seventy-nine urban young adults (18–25 years) provided matched pre-lockdown and during-lockdown data via an online survey. Measures included the Pittsburgh Sleep Quality Index (PSQI), Berlin Questionnaire for OSA risk, the National Stressful Events Survey Acute Stress Disorder Short Scale (NSESS-S), and self-reported height and weight for BMI. Social jet lag was derived from sleep timing. Within-participant changes were tested using Wilcoxon signed-rank tests; univariate and multivariate logistic regressions identified predictors of OSA risk.

No significant changes were observed in PSQI scores, subjective sleep quality, sleep latency, acute stress, or social jet lag during lockdown. Sleep duration increased slightly, and BMI and social jet lag showed small numerical rises without statistical significance. OSA risk was positively associated with acute stress (p < 0.001), higher BMI (p < 0.05), and poor sleep quality (p < 0.001). In multivariate models, poor sleep quality was the strongest independent predictor of OSA risk, followed by acute stress and BMI. Lifestyle factors, including physical activity, screen time, and ambient noise, showed no significant associations.

Flexible schedules during lockdown may have offset expected negative impacts on sleep and stress in this demographic. The strong links between OSA risk, poor sleep quality, and acute stress highlight the need for integrated behavioural and physiological approaches to sleep health in young adults.

## INTRODUCTION

Stress and sleep quality are closely linked, forming a bidirectional relationship in which one can act synergistically with the other^1,2^. Heightened psychological stress can delay sleep onset, cause fragment sleep, and reduce sleep efficiency^3^. In turn, curtailed or poor quality sleep increases next day stress reactivity^4^. This interaction has important public health implications, as even short periods of disturbed sleep can negatively influence cognition, emotional stability, and cardiometabolic regulation^4,5^.

Young adults represent a particularly vulnerable group within this framework. Before the COVID-19 pandemic, university and early-career populations consistently showed a high prevalence of poor sleep quality^3,6^. Academic demands, irregular schedules, and the transition toward greater independence coincided with elevated perceived stress, making this age range susceptible to disturbances in routine that can disrupt sleep wake patterns^7–9^.

The onset of the pandemic intensified these vulnerabilities several folds. Large scale lockdowns, abrupt shift to remote learning and work, and restrictions on in-person social interaction altered exposure to environmental time cues, increased screen use, and reduced opportunities for physical activity^10,11^. International reports suggest that while the flexibility of lockdown sometimes allowed for longer time in bed, subjective sleep quality did not consistently improve, and variability in sleep timing remained common^12,13^. These mixed outcomes underline the need to consider timing, duration, and perceived quality of sleep as well, rather than duration alone^14^.

Circadian alignment plays a key role in interpreting these changes, as reduced exposure to zeitgebers like morning light can impair synchronization of biological rhythms^15^. Social jet lag, defined as the discrepancy between the midpoint of sleep on workdays and free days, serves as a measure of misalignment between biological rhythms and social schedules^16,17^. The removal of fixed early start times during lockdown could theoretically reduce this misalignment; however, irregular daily demands, increased evening screen exposure, and reduced structure may offset such benefits^18^. At the same time, the pandemic environment fostered conditions that increased acute stress symptoms, well known to impair sleep continuity and depth^17,19^.

Despite the global nature of these disruptions, data from India remain limited^14,20^, especially from studies that compare the same individuals before and during lockdown. Furthermore, few investigations have examined multiple interrelated variables such as sleep quality, sleep duration, social jet lag, acute stress, and body mass index (BMI) in young adults, despite the likelihood that each can shift under confinement and influence the others.

The present study evaluated sleep and stress of Indian young adults using a within-participant approach, assessing data from both before and during the lockdown period. We measured sleep quality, sleep duration, and social jet lag alongside self-reported acute stress and BMI. We further examined how acute stress and social jet lag related to sleep quality under pandemic conditions. The aim was to generate context specific evidence on how lockdown conditions affected sleep and stress, and to clarify the interrelationship between circadian misalignment, psychological stress, and subjective sleep quality during a period of profound routine disruption.

## MATERIALS AND METHODS

### Study design

This observational study was conducted through an online survey administered to urban Indian youth during the COVID-19 pandemic. Participant recruitment flowchart is shown in Figure 1. Ethical clearance was obtained from the Institutional Ethics Committee (IECPG-654/28.10.2021, RT-01/27.01.2022), and informed consent was taken from all participants prior to their inclusion in the study. A total of 502 volunteers completed the questionnaire, of whom 130 provided data both before and during the lockdown. After applying the inclusion and exclusion criteria, 79 participants were included in the final analysis. Eligible participants were between 18 and 25 years of age and residing in urban areas. Individuals undergoing treatment for sleep disorders or outside the specified age range were excluded.

**Figure 1:**
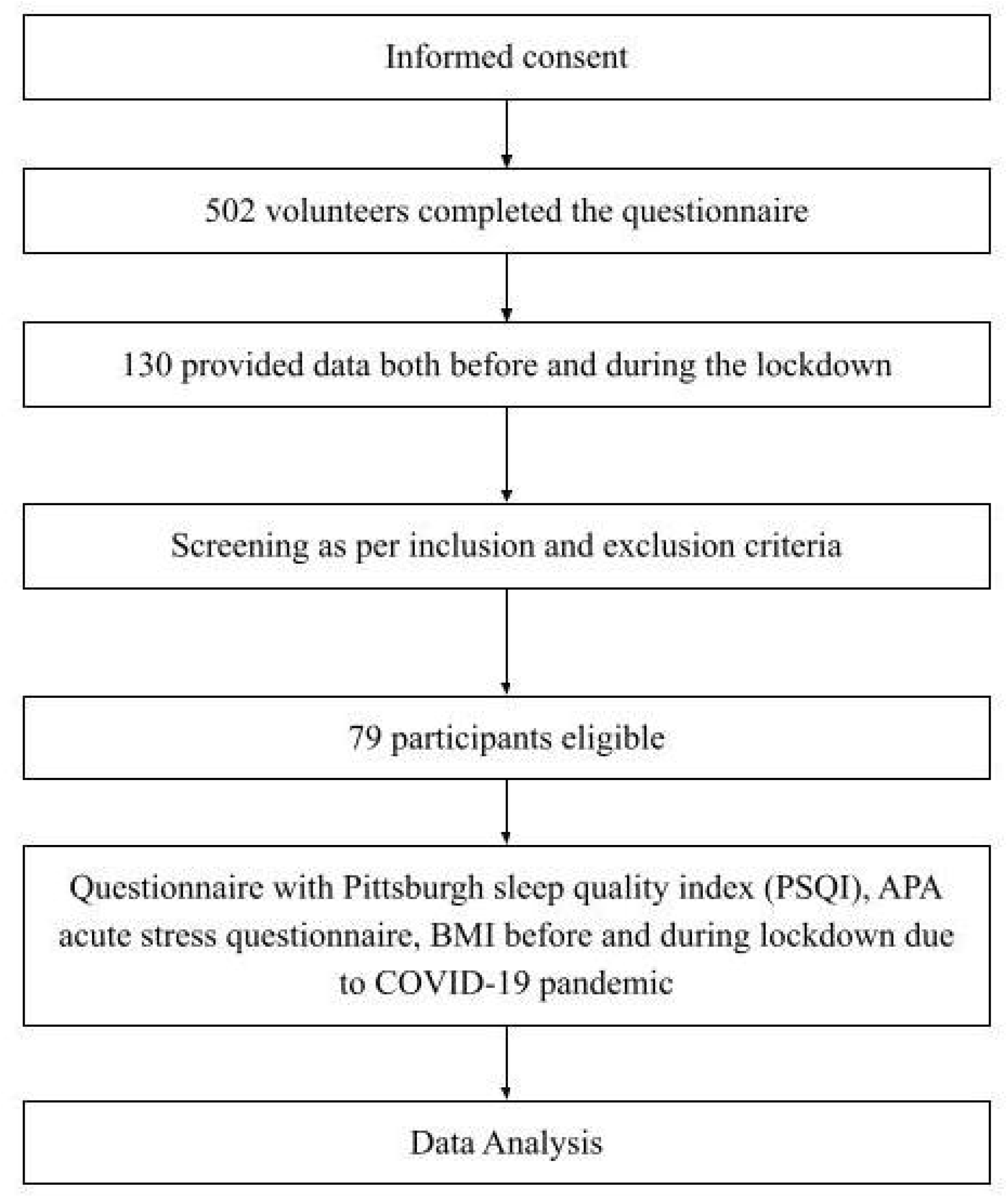
Participant recruitment flow diagram.

### Data collection and measures

Participants completed a comprehensive online questionnaire that included the Pittsburgh Sleep Quality Index (PSQI)^21^ to assess sleep quality, the Berlin Questionnaire for sleep apnoea risk^22^, the APA’s National Stressful Events Survey Acute Stress Disorder Short Scale (NSESS-S) for acute stress assessment^23^, and self-reported height and weight to calculate BMI. Sleep duration and midpoints on workdays and free days were also collected to derive social jet lag.

Social jet lag was calculated using the corrected midpoint of sleep on free days (MSFsc), following the method proposed in a previous study^24^:

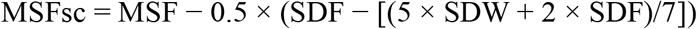

where **MSF** = midpoint of sleep on free days, **SDF** = sleep duration on free days, and **SDW** = sleep duration on workdays

Social jet lag (SJL) was then calculated as:

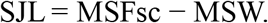

### Sample size calculation

A post-hoc power analysis was performed in G*Power (version 3.1.9.7, Universität Düsseldorf, Germany)^25^ for a paired-samples t-test, using α = 0.05 (one-tailed) and the final sample size of 79. The effect size, calculated from pre- and post-COVID mean values of social jet lag, was Cohen’s dz ≈ 0.56. The analysis indicated that the achieved power to detect a within-participant change of this magnitude was 0.9995, corresponding to a medium effect size. This suggests that the sample size was more than adequate to detect meaningful changes in this outcome.

### Statistical analysis

Statistical analysis was performed using GraphPad Prism version 9.4 (GraphPad Software, Boston, Massachusetts USA, www.graphpad.com). The Wilcoxon signed-rank test was used for within-subject comparisons of non-parametric data. Univariate and multivariate logistic regression analyses were conducted to identify associations between sleep apnoea risk and variables such as sleep quality, stress, BMI, and social jet lag.

## Results

### Participant characteristics

Of the 502 volunteers who completed the survey, 130 participants submitted responses for both the pre-lockdown and during-lockdown phases. After applying the eligibility criteria, data from 79 participants were included in the final analysis. The mean age of participants was 22.7 ± 2.7 years, with 43 males and 36 females. The mean BMI increased from 23.44 ± 4.63 kg/m^2^ before the COVID-19 lockdown to 24.81 ± 6.14 kg/m^2^ during the lockdown (Table 1).

**Table 1:** Demographic characteristics of participants.

| Parameter | Value |
| --- | --- |
| Age (years) | $22.7 \pm 2.7$ |
| Gender (M/F) | 43/36 |
| BMI pre COVID (kg/m<sup>2</sup>) | $23.44 \pm 4.63$ |
| BMI post COVID (kg/m<sup>2</sup>) | $24.81 \pm 6.14$ |

### Changes in sleep, stress, and social jet lag

Comparison between pre lockdown and during lockdown data showed no statistically significant changes in PSQI scores, sleep latency, acute stress scores, subjective sleep quality, or social jet lag (Figures 2 and 4). Mean sleep latency increased slightly, PSQI scores suggested marginally better sleep quality, and acute stress scores also increased; however, none of these changes reached statistical significance. Sleep duration showed a numerical increase, and social jet lag increased slightly during lockdown, but neither change was statistically significant.

**Figure 2:**
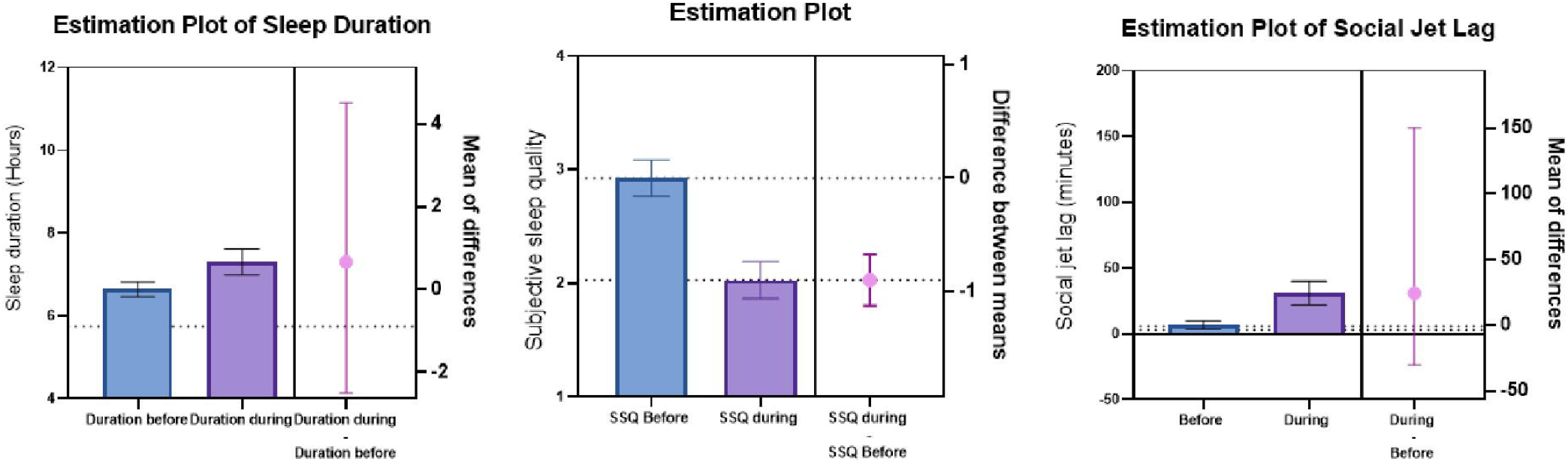
Changes in sleep duration, subjective sleep quality, and social jet lag in young adults before and during the COVID-19 lockdown. Estimation plots showing within-subject changes in sleep duration, subjective sleep quality, and social jet lag. Bars represent mean ± SEM; paired differences are displayed with 95% confidence intervals. Statistical significance was assessed using Wilcoxon signed-rank tests.

### Associations of sleep apnoea risk with psychological and physiological Factors

Risk of sleep apnoea showed a significant positive association with acute stress (p < 0.001), BMI (p < 0.05), and poor sleep quality (p < 0.001) (Figure 3). However, no significant associations were found between sleep apnoea risk and ambient noise levels, screen time, physical activity, or social jet lag.

**Figure 3:**
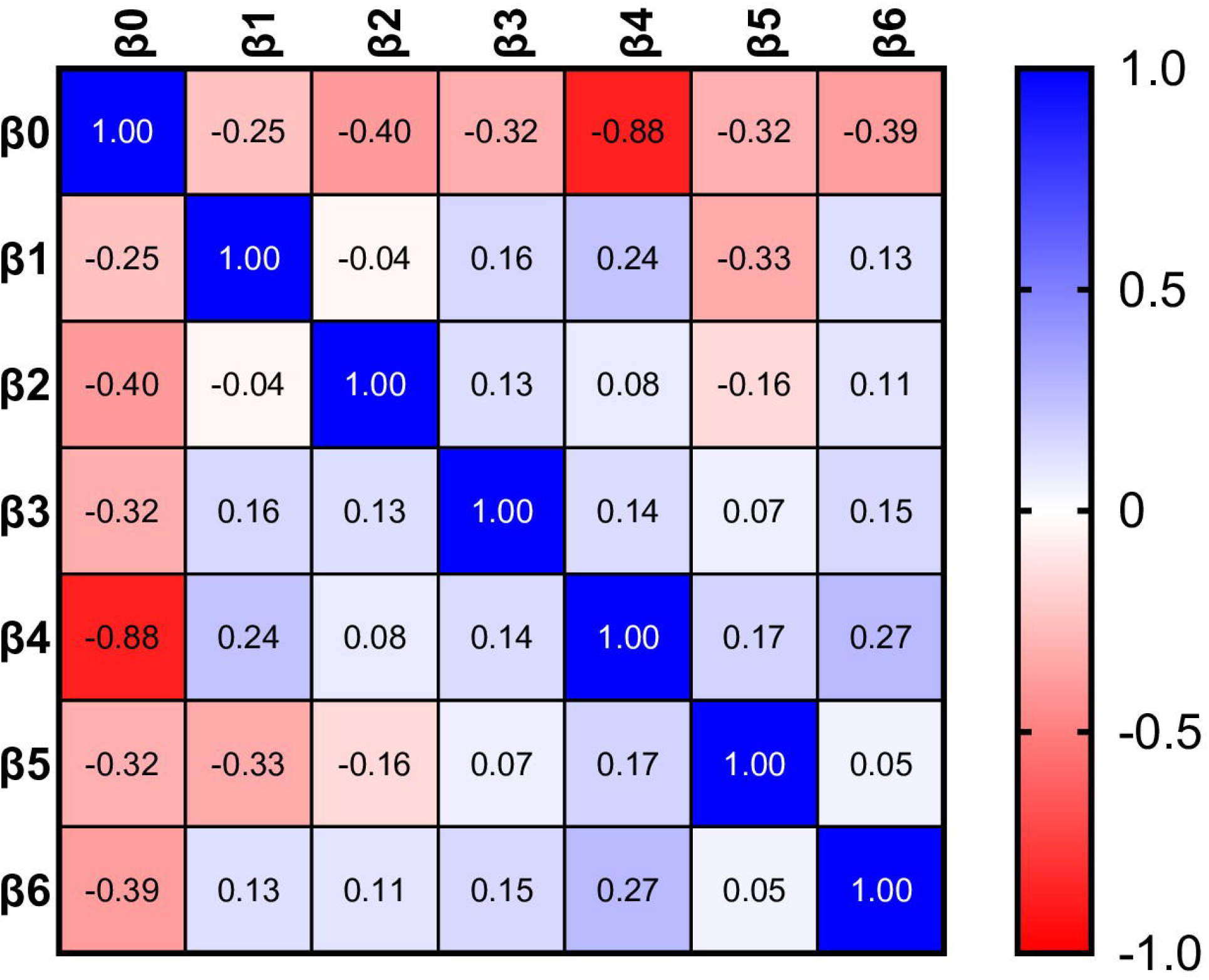
Correlation matrix of predictor variables in the regression model. Heatmap displaying Pearson correlation coefficients between predictor variables (β[–β[) used in the regression model. The color scale ranges from –1 (strong negative correlation, red) to +1 (strong positive correlation, blue), with white indicating no correlation. Strongest negative correlation was observed between β[and β[(r = –0.88).

**Figure 4:**
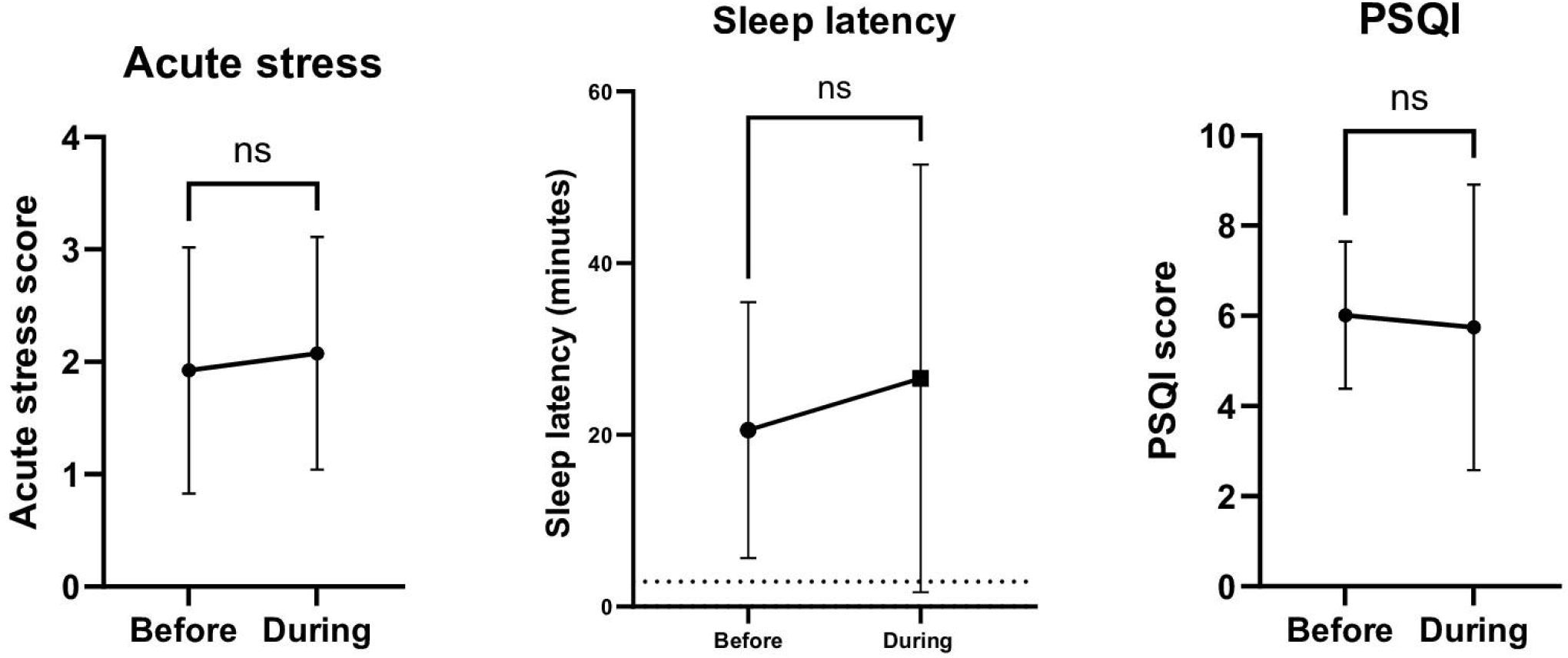
Changes in sleep latency, PSQI score, and acute stress before and during the COVID-19 lockdown. Mean sleep latency increased, PSQI score suggested better sleep quality, and acute stress scores increased; however, none of these changes reached statistical significance (ns). Data are presented as mean ± SD; comparisons were made using Wilcoxon signed-rank tests.

### Regression analysis of predictors of sleep apnoea risk

In univariate logistic regression analysis, the odds of having a high risk of sleep apnoea increased significantly with the severity of acute stress, with individuals experiencing extreme acute stress having nearly 15 times higher odds. However, social jet lag did not show a significant association in this model. Multivariate regression revealed that poor sleep quality had the strongest independent association with sleep apnoea risk, followed by acute stress and being overweight (Table 2).

**Table 2:** Multivariate regression analysis for predictors of subjective sleep quality after the COVID-19 lockdown. Parameter estimates, standard errors, 95% confidence intervals, Z-scores, and p-values for variables included in the multivariate regression model with subjective sleep quality as the dependent variable. Predictors included sleep latency, wake-up time, sleeping time, sleep duration, acute stress, and social jet lag measured during the lockdown period. None of the predictors were significantly associated with subjective sleep quality (all p > 0.05).

| Parameter estimates | Variable | Estimate | Standard error | 95% CI (profile likelihood) | \|Z\| | P value | P value |
| --- | --- | --- | --- | --- | --- | --- | --- |
| $\beta_0$ | Intercept | 0.6738 | 0.6189 | -0.5563 to 1.870 | 1.089 | 0.2763 | ns |
| $\beta_1$ | Sleep Latency after COVID | 0.004263 | 0.003182 | -0.002302 to 0.01020 | 1.340 | 0.1803 | ns |
| $\beta_2$ | Wake up time after COVID | 0.01232 | 0.02427 | -0.03702 to 0.05827 | 0.5078 | 0.6116 | ns |
| $\beta_3$ | Sleeping time after COVID | -0.0001149 | 0.01086 | -0.02203 to 0.02066 | 0.01058 | 0.9916 | ns |
| $\beta_4$ | Sleep duration during lockdown | -0.04476 | 0.06192 | -0.1664 to 0.07648 | 0.7228 | 0.4698 | ns |
| $\beta_5$ | Acute stress after COVID | 0.06732 | 0.08166 | -0.09402 to 0.2265 | 0.8244 | 0.4097 | ns |
| $\beta_6$ | Social jet lag after COVID | -0.0007642 | 0.002106 | -0.005004 to 0.003267 | 0.3629 | 0.7167 | ns |

## Discussion

The present study investigated the impact of the COVID-19 lockdown on sleep characteristics, acute stress, and social jet lag in Indian young adults, alongside their associations with risk of obstructive sleep apnoea (OSA). Contrary to concerns that the disruption of daily schedules during lockdown would impair sleep, our data showed no statistically significant changes in PSQI scores, subjective sleep quality, sleep latency, acute stress scores, or social jet lag. Sleep duration increased slightly during lockdown, consistent with reports from other populations where reduced commuting time and flexible schedules permitted later wake times and extended rest periods^26^. These findings suggest that the lockdown period did not produce large-scale deterioration in sleep or stress indices in this cohort.

From a physiological standpoint, the absence of significant deterioration in sleep quality may be explained by the reduction in early morning social constraints, a factor known to reduce weekday–weekend misalignment and improve alignment between endogenous circadian rhythms and behavioural schedules^27^. However, the slight increase in social jet lag observed suggests that not all participants adopted consistent bedtimes and wake times, potentially due to irregular academic demands or increased late-night screen exposure, both of which can delay circadian phase through suppression of evening melatonin secretion^26,28^. The absence of significant changes in acute stress could indicate resilience within this age group or that stressors specific to the pandemic were balanced by the removal of other daily pressures, such as commuting and in-person academic obligations.

The association analyses identified links between sleep apnoea risk and acute stress, BMI, and poor sleep quality. OSA is characterised by recurrent episodes of upper airway collapse during sleep, leading to intermittent hypoxia, sleep fragmentation, and consequent activation of the hypothalamic–pituitary–adrenal (HPA) axis^29^. Elevated acute stress can exacerbate airway collapsibility through increased sympathetic drive and altered upper airway muscle tone^28,30^, while poor sleep quality, whether due to OSA itself or other factors, may further potentiate stress reactivity. The positive association with BMI aligns with increased adiposity, particularly central fat deposition, narrows the upper airway and heightens its collapsibility^31^. The regression analyses suggest sleep quality as an independent predictor of OSA risk in this cohort, even after adjusting for BMI and stress. The strong link with acute stress also highlights the need to consider psychophysiological interactions in OSA screening, especially in populations exposed to prolonged environmental or situational stressors.

Our study had few limitations which should be acknowledged. Our study did not find significant relationships between OSA risk and lifestyle factors, might be because these exposures were self-reported and may have lacked the sensitivity to detect smaller effects. Similarly, social jet lag did not emerge as a significant predictor of OSA risk, which may reflect the relatively small variability in workday versus free day schedules during lockdown when most participants were engaged in remote learning. In our study, sleep quality, sleep timing, stress, and lifestyle factors were assessed using self-report questionnaires, which are subject to recall bias and may not capture objective physiological changes. The PSQI and other tools capture a one-month recall window, which may dilute the detection of transient changes. Also, the cross-sectional nature of the association analyses limits causal inference between OSA risk and its correlates. Finally, the study was conducted in urban populations, and results may not generalise to rural or socioeconomically diverse settings where lockdown experiences differed substantially.

A key strength of this study is its within participant design, comparing the same individuals before and during lockdown, which controls for inter-individual variability in baseline sleep characteristics and stress responses. The inclusion of multiple interrelated variables such as sleep quality, sleep duration, social jet lag, acute stress, BMI, and OSA risk, provides a multidimensional view of sleep, stress, and health interactions in this demographic. The use of validated instruments such as the PSQI, Berlin Questionnaire, and NSESS-S enhances the reliability and comparability of findings with existing literature.

Future research should incorporate objective measures of sleep and circadian timing, such as actigraphy or polysomnography, to validate and extend self-reported data. Given the association between acute stress and OSA risk, longitudinal designs could clarify whether heightened stress during prolonged environmental disruptions predisposes to incident OSA. Further, expanding research to rural and peri-urban populations could reveal whether differing lockdown conditions and socioeconomic contexts modify these relationships. Finally, mechanistic studies exploring the role of autonomic function, inflammatory markers, and hormonal stress responses could help elucidate the physiological pathways linking stress, BMI, and OSA risk in young adults.

## Conclusion

As compared to before the pandemic, during the lockdown young adults had a better subjective sleep quality, increased sleep duration, Increased BMI and increased social jet lag. There was no change in sleep latency, wake up time, time for onset of sleep and acute stress level.

## Data Availability

Available upon reasonable request to corresponding author.

